# AVATAR therapy for distressing auditory hallucinations: mediators of change in AVATAR1, a single blind randomised control trial with supportive counselling as active control

**DOI:** 10.64898/2026.08.11.26360172

**Authors:** Miriam Fornells-Ambrojo, Anca Chis Ster, Philippa Garety, Tom K J Craig, Mark Huckvale, Richard Emsley, Clementine Edwards, Amy Hardy, Thomas Ward, Mar Rus Calafell

## Abstract

AVATAR therapy is an effective relational therapy for persistent distressing auditory verbal hallucinations (voices). A digital representation of the embodied persecutory voice (avatar) is created and used in a series of dialogues in which the voice hearer is supported to be more assertive and the avatar concedes power. In the first mediation analysis of AVATAR therapy examining the role of power-related constructs, we investigate whether treatment effects on total severity, frequency, and distress of voices are mediated by changes in beliefs about voices and the self, voice relationship appraisals and anxiety. Mediation effects were evaluated in relation to decomposing treatment offer and treatment receipt effects using both Intention to treat (ITT) and Complier Average Causal Effect (CACE) analyses. One hundred and fifty participants from AVATAR1, a randomised control trial (RCT) comparing AVATAR therapy to Supportive Counselling took part in this study, with their baseline and end of treatment (12 weeks) data used. As hypothesised, across both ITT and CACE analyses, reductions in perceived voice omnipotence and increased assertiveness in relation to voices emerged as consistent mediators of AVATAR therapy on reductions in overall severity, frequency and distress of auditory hallucinations compared to SC, whereas voice malevolence, perceived power differential, self-esteem and anxiety did not. Exploratory analysis also indicated that increases in acceptance and autonomy in relation to voices mediated the impact of AVATAR therapy on overall voice severity and distress. This mediation analysis refines our understanding of AVATAR therapy and highlights agency, voice omnipotence and acceptance as intervention targets.

## Introduction

Auditory verbal hallucinations (AVH), also referred to as voices, are often distressing experiences among people with psychosis requiring clinical care [1, 2]. Antipsychotic medication has moderate efficacy, with 20-35% reporting no clinically significant improvements [3-5]. Such voices are often dominant, persecutory in nature, making threats, criticising and abusing the voice hearer, in a way that reflects developmental interpersonal trauma and ongoing social adversity [6-10]. The often-devastating impact on the voice hearer’s life has been understood in the context of beliefs about power and control, sense of self in cognitive models of voices [11] and subordination as an evolutionary survival tool in social rank theory [12]. Additionally, experiential avoidance and cognitive fusion are highlighted by Acceptance and Commitment therapy (ACT) [13], and the intrusiveness of threatening memories associated with fear responses and anxiety in traumatogenic views of voice hearing^1^ [14-16].

The current gold standard psychological therapy for psychosis, Cognitive Behavioural Therapy for psychosis (CBTp) which aims to reduce voice related distress by modifying voice appraisals, has shown small to medium effect sizes [17]. More recently, a new wave of relational therapies for voices [18] that use experiential dialogue to engage directly with voices or indirectly with identities associated with them as their core therapeutic intervention are taking centre stage. Their common focus is on patterns of interaction with voices and relational dynamics that include, but go beyond, dominance and submissiveness. Amongst these relational therapies, AVATAR therapy has, to date, been the more extensively researched internationally by fully powered trials, showing evidence of efficacy. However, its mechanisms of change are still poorly understood.

### What is AVATAR therapy and how does it work?

AVATAR therapy uses digital technology to create a digital representation of the embodied persecutory voice (i.e. the avatar). The software enables the voice-hearer to personalise their avatar to match their lived experience of voice hearing. Therapy involves engaging dialogically with the avatar with the support of the therapist, aiming to achieve therapeutical. AVATAR therapy as delivered in Craig et al., 2018 is described as comprising two phases, phase one focussing on encouraging the voice hearer to be more assertive to the dominant voice while phase two begins with the avatar conceding power, becoming more dialogic and adopting a conciliatory position [19, 20]. Meta-analysis of eight trials (n=978) revealed that AVATAR therapy resulted in moderate reductions in AVH severity (Hedges’g =−0.40,95%CI−0.54 to−0.25), and small reductions in voice distress (Hedges’g = −0.32, 95% CI −0.46 to −0.18), and frequency (Hedges’g = −0.38, 95% CI −0.52to −0.24) compared with control conditions [21].

For complex interventions such as AVATAR therapy, mechanism evaluations are recommended to understand the process through which the intervention influences outcomes [22, 23]. These mechanistic processes are commonly referred to as mediators. Mediation analysis seeks to decompose an overall treatment effect into a mediated effect, operating through a specified mediator, and a direct effect, capturing effects operating through all other pathways. When the primary analysis of a clinical trial estimates the intention-to-treat (ITT) effect based on randomisation group, the total ITT effect may be decomposed into a Natural Direct Effect (NDE) and a Natural Indirect Effect (NIE) under appropriate identification assumptions [24].

No formal mediation analysis has examined at the role of power-related constructs in AVATAR therapy, but existing qualitative studies provide lived experience insights into the processes by which the therapy may lead to change [25]. This work suggests that voice hearers who receive AVATAR therapy [26-28] feel empowered to stand up to the avatar and experience increases in power and control, even though the dialogues are anxiety provoking. These new ways of relating to voices are also reported to positively impact everyday social encounters.

Consistent with these subjective reports, fine grained analysis of avatar dialogues revealed nuanced communication around relational power, with early assertiveness work reinforced by the therapist, and voice hearer’s submissive behaviours reducing gradually during therapy [29]. Whilst Glenthøj and colleagues found that generic emotion regulation strategies do not mediate the impact of AVATAR therapy on voice severity [30], the specific mechanisms through which AVATAR therapy achieves change, particularly shifts in voice-related beliefs and interpersonal responding, have yet to be formally examined. Several converging lines of evidence point towards these as plausible candidates. First, although high sense of voice presence (i.e. the subjective experience of talking to the voice) did not in itself predict reductions in voice frequency [31, 32], its interaction with anxiety reductions was associated to positive outcomes. This suggests that in order to achieve change realism during avatar exposure is crucial in triggering fear responses when talking to the avatar [31, 32]. Second, experience sampling data indicate that momentary anxiety is reduced following AVATAR therapy, but only when the voice is thought about rather than actively heard [33], suggesting that change operates through anticipatory appraisals of the voice rather than direct exposure alone. Together, these findings point to belief change and relational shifts as the core active ingredients of AVATAR therapy,

### The current study

AVATAR1^2^, the first fully powered RCT comparing AVATAR therapy to supportive counselling (SC) set out to investigate such explanatory mechanisms of action [34] with SC serving as a control for non-specific factors, such as an empathic, therapeutic relationship that what is known to impact outcomes [26, 35]. This study showed large effect sizes (d=0.8) on the prespecified primary outcome of overall severity of voices, but also significant improvements in voice frequency and distress [36]. The perceived omnipotence of voices reduced also at the end of treatment and voices were more likely to be accepted and less likely to be acted upon. Unexpectedly, however, voice malevolence, relative power, anxiety and self-esteem did not significantly differ between treatment groups.

The current paper reports the findings of a planned mediation analysis of both ITT effects and response to treatment adherence. ITT mediation analyses evaluate the mechanisms of treatment offer rather than actual treatment exposure — that is, the processes through which clinical benefit is hypothesised to be achieved. To address how the treatment works when received, the total Complier Average Causal Effect (CACE) analyses were conducted, which estimate treatment efficacy specifically among those who engaged with the intervention. The CACE can then be decomposed into a mediated effect, the Complier Average Causal Mediated Effect (CACME), and a direct component, the Complier Average Natural Direct Effect (CANDE) [37, 38] under a set of extended instrumental variables (IV) assumptions.

### Aims and hypotheses

The aims of the study are to:

1. Test the mechanisms of change hypothesised in the AVATAR1 trial protocol. Specifically, that the mediators of treatment effects of AVATAR therapy on changes in auditory hallucinations (overall severity, frequency and distress) will be:

- Beliefs about voices (specifically reductions in omnipotence and malevolence)
- Beliefs about the self (improved self-esteem)
- Appraisal of voice relationship (specifically increase in relative power compared to the avatar and increased assertiveness)
- Reductions in anxiety
2. Explore additional mediators (voice acceptance and action) that were shown to significantly improve with AVATAR therapy [36].
3. Both aims will be addressed using mediation analyses conducted within an Intention-to-Treat (ITT) framework and a Complier Average Causal Effect (CACE) framework, allowing the effect of treatment offer to be distinguished from treatment receipt on these mechanisms.

## Methods

### Trial design

This was a single-blind randomised controlled trial at a single location (South London and Maudsley NHS Foundation Trust) (ISRCTN65314790), comparing AVATAR therapy to SC. The trial received ethical approved from the London-Hampstead Research Ethics Committee (reference 13/Lo/0482). Full details on the design, demographics and trial findings can be found in the trial protocol [34] and main trial results publication [36]. The current mediation study includes outcomes and potential mediators measured at end of treatment (12 weeks) by blinded assessors. This time point was selected because it is the primary outcome period of the study [36].

### Participants

One hundred and fifty participants consented to the trial, with n=75 allocated to AVATAR therapy and n=75 to SC. Key inclusion criteria were presence of enduring auditory verbal hallucinations during the previous 12 months, despite continued psychopharmacological treatment, being aged 18 to 65 years, with a clinical diagnosis of a schizophrenia spectrum (ICD10 F20–29) or affective disorder (F30–39 with psychotic symptoms). Please see further details in the trial protocol [34].

### AVATAR therapy intervention and supportive counselling (SC)

After completing the set-up of the avatar in an introductory session, which includes a comprehensive assessment of the voice(s), the therapy was delivered over typically six (but up to 9) weekly 50-min sessions. Phase 1 (typically sessions 1 to 3) involves *exposure* and *assertiveness*, with the avatar initially voicing verbatim voice content (i.e. abusive, threatening, critical) and transitioning to a more conciliatory position towards the end of this phase. In phase 2 (typically sessions 4-6) the therapist continues addressing power and autonomy resulting in a gradual reduction of voice hearers’ submissive behaviours, with the avatar-voice hearer dialogue addressing developmental and relational understanding of voices [20, 29, 36]. SC, the active control condition, was delivered over six sessions [39, 40]. In practice, the average number of sessions attended was 5·6 (SD 2·8, range 0–10) for AVATAR therapy and 5·1 (3·1, 0–10) for SC [36].

### Definition of adherence

Number of AVATAR sessions attended was chosen as a proxy for treatment adherence. Completers were defined as those who completed the introductory session (set-up of the avatar) as well as at least three of the six active (dialogue) AVATAR therapy sessions, and non-adherers those who attended two or less. This threshold was based on therapeutic phases of AVATAR therapy so that adherers had completed Phase I (exposure to voice content with the therapist encouraging assertive responding. A sensitivity analysis will define adherers as having attended at least one session of the six active AVATAR therapy sessions to assess the robustness of the exclusion restriction assumptions.

### Measures

Please see the main AVATAR therapy protocol and outcome paper [34, 36] for further details on measures.

#### Outcomes

The prespecified primary outcome was the total score (0–44) on the Psychotic Symptom Rating Scales, auditory hallucinations subscale (PSYRATS–AH) [41]. The secondary outcome measures of voices were dimensional subscales of the PSYRATS-AH [42]: voice frequency (frequency, duration, and disruption items) and voice distress (negative content, distress, and control items).

#### Mediators

Eight potential mediators grouped into four conceptual domains were examined.

##### Beliefs about voice omnipotence and malevolence

The first two hypothesised mediators of AVATAR therapy were assessed by two subscales of Beliefs About Voices Questionnaire–Revised (BAVQ-R) [43]. *Omnipotence*, assessed with 6 items covering absolute voice power and dominance (e.g. ‘my voice is very powerful’, ‘the voice rules my life’), a sense of omniscience (e.g. ‘the voice seems to know everything about me’) as well as feared consequences of disobedience (e.g. ‘My voice will harm or kill me if I disobey or resist it’). The second subscale, *malevolence,* includes 6 items that capture the perceived harmful intentions of the voice (e.g. ‘my voice wants to destroy me’). Higher omnipotence and malevolence, where the voice is believed to be more powerful and with more malign intentions, are associated with poorer outcomes such as depression [44].

##### Relative power and assertiveness

The third and fourth mediators drawn from the Voice Power Differential Scale (VPDS) scale [45], were the *power differential* between the voice and the voice hearer (VPDS1) where higher scores indicate high power differential (subordination) *(i.e. ‘* I am much more powerful than my voice’ [1] –‘My voice is much more powerful than me’ [5]) assessed *on a* 5-point-Likert-scale, and an item on *assertiveness* (VPDS8) added for the purposes of AVATAR1 [34], which measures the voice hearer’s ability to assert their own needs to the voice (i.e. ‘I can always tell the voice what I want’). Lower scores indicate higher assertiveness.

##### Voice acceptance and autonomous action

Voices Acceptance and Action Scale (VAAS) [46] contains two subscales: *acceptance* of the voice hearing experience (e.g. ‘I have learned to live with my voices’) and *action,* which assesses autonomous actions towards voices (e.g. ‘Just because a voice tells me to do something, it doesn’t mean I have to do it’).

##### Self-esteem and anxiety

Non-specific voice hypothesised mediators were *self-esteem,* assessed by the Rosenberg Self-Esteem Scale (RSES) [47] and *anxiety*, derived from the Depression Anxiety and Stress Scale 21 (DASS-21) [48].

### Statistical analysis

This study is reported according to ‘A Guideline for Reporting Mediation Analyses’ (AGReMA) [49]. Please see Supplementary Materials.

#### ITT analysis in the main outcomes paper

As reported in the main outcomes paper [36] the primary analysis model used the original randomised groups (Intention-to-treat ITT analysis), with a linear mixed-effects model with baseline measurement of PSYRATS-AH and randomisation as fixed effects, and a random intercept for each therapist in the two groups to allow for differential therapist clustering by randomised group. As hypothesised, the reduction in PSYRATS–AH total score at 12 weeks was significantly greater for AVATAR therapy than for SC (mean difference −3·82 [SE 1·47], 95% CI −6·70 to −0·94; p<0·0093). There were also significant differences in reported frequency of voices (mean difference −1·22 [SE 0.38], 95% CI −1.97 to −0.48; p=0·0013) and reduced distress (mean difference −2.34 [SE 0.98], 95% CI −4.26 to −0.42; p=0·017) at 12 weeks.

#### Mediation analysis of the ITT effect

We decomposed the total ITT effect for each mediator into a Natural Indirect Effect (NIE) and a Natural Direct Effect (NDE). For identification, we assume that SUTVA, positivity, conditional treatment ignorability, and sequential ignorability assumption all hold [50]. For each outcome and each mediator, the ITT, NIE, and NDE are estimated using linear Structural Equation Models (SEMs) as depicted in Error! Reference source not found.**a.**[51] The NDE then corresponds to path *c’* in Error! Reference source not found.**a**, and since all measures are continuous the NIE corresponds to the product of the paths *a* and *b* and the (total) ITT is the sum of the NIE and NDE estimates.

#### Mediation analysis of CACE

We decomposed the CACE into a CACME and CANDE [37] for each mediator and outcome. For identification, we assume treatment relevance, conditional treatment ignorability, monotonicity (no defiers), exclusion restrictions for the mediator and outcome for noncompliers (i.e., randomisation can influence the mediator and outcome only through receiving treatment), no unmeasured mediator-outcome confounding among compliers, and no interaction between treatment receipt and the mediator on the outcome. The instrument (randomisation) for treatment receipt implicitly handles any confounding in the treatment receipt – mediator and treatment receipt – outcome relationships. We estimated the CACE, CACME, and CANDE with linear SEMs as depicted in **Figure 1b**. The estimation approach assumes (i) linearity for any continuous covariates in the model for the mediator (ii) a linear effect of the continuous mediator on continuous outcome (iii) no interaction between treatment receipt and mediator on the outcome and (iv) linearity for any other continuous covariates in the model for Y. [51]The CANDE then corresponds to path *C’* in **Figure 1b**, the CACME corresponds to the product of the paths *A* and B, and the (total) CACE is the sum of the CANDE and CACME.

**Figure 1.**
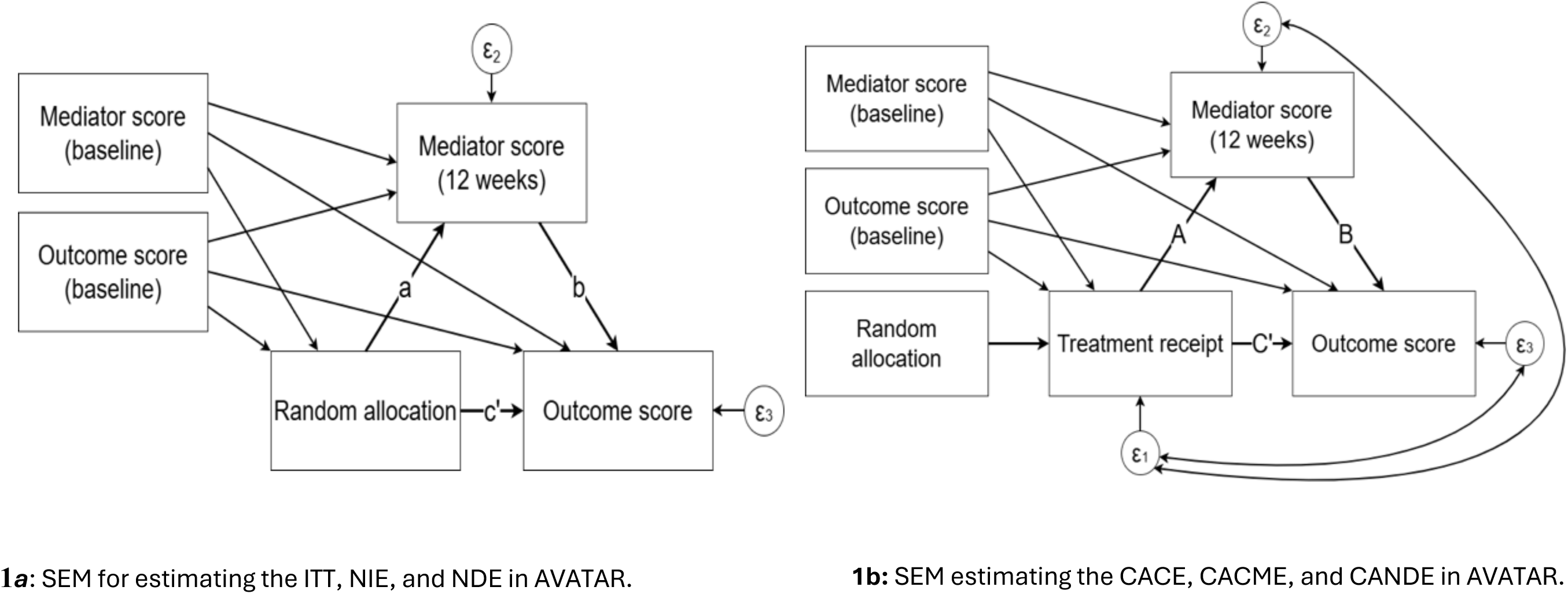
Structural equation models (SEMS) ***Note:*** *The SEM figure does not explicitly depict the multilevel structure for clustering by therapist.* Error variances for treatment receipt, mediator, and outcome, are represented by the variables *ɛ*_1_, *ɛ*_2_ and *ɛ*_3_, respectively. **Abbreviations:** Intention-to-treat (ITT); natural indirect effect (NIE); natural direct effect (NDE); complier average causal effect (CACE); complier average causal mediated effect (CACME); complier average natural direct effect (CANDE); standard error (SE); confidence interval (CI)

In both mediation analyses of the ITT and CACE, baseline mediator and outcome scores were included as covariates in the analysis models for increased precision [51] and to improve plausibility of the no mediator-outcome confounding assumptions. Cluster-robust standard errors (SEs) were implemented to account for the clustering by therapist, and CIs were estimated using nonparametric percentile bootstrap with 1,000 replications. Analyses were computed with Stata SEM software [52]. The Stata SEM program handles missing values under a complete case analysis and implicitly assumes data are missing at random (MAR) [53, 54].

## Results

Table ***1*** presents summary statistics for the outcomes and the potential mediators at baseline and 12 weeks. Of the 150 individuals, 37 missing baseline scores for VPDS 1 *power* and VPDS 8 ￼[46]￼, and 21 missing baseline scores for *anxiety* (￼[48]￼ were mean imputed.

**Table 1:**
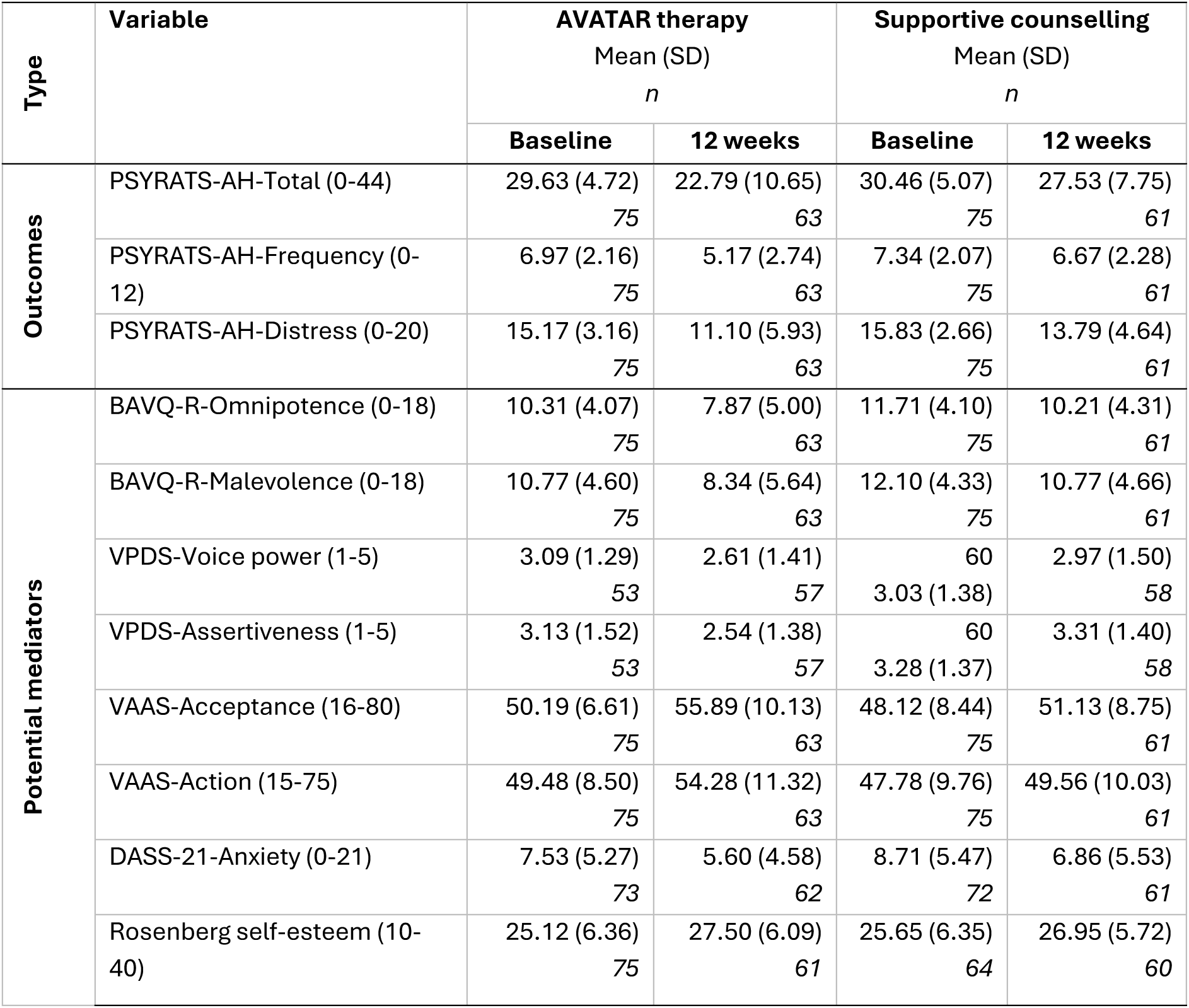
Means and standard deviations for outcomes and potential mediators and at baseline and 12 weeks

Error! Reference source not found. shows the ITT and CACE mediation analyses for the overall severity of voices PSYRATS-AH-Total outcome at 12 weeks, along with bootstrap SEs and 95% percentile bootstrap confidence intervals (CIs).

### ITT mediation

The potential mechanisms of the effect of random allocation to AVATAR therapy on PSYRATS-AH-Total score at week 12 were *omnipotence*, *assertiveness*, voice *acceptance* and *autonomous actions*. For example, as can be seen for the voice omnipotence mediator, adjusting for baseline *omnipotence* and PSYRATS-AH-Total score, an ITT analysis suggests that the average causal effect of offering AVATAR therapy decreases PSYRATS-AH-Total score at week 12 by 3.68 points (SE = 1.59, 95% CI -6.73 to -0.70). AVATAR treatment offer decreased the overall severity of voices (PSYRAYS-AH-Total score) at week 12 via a reduction in the participants perceived *omnipotence* of the voice by 1.37points (SE = 0.91, 95% CI -3.55 to -0.04), while holding all other pathways constant. The corresponding natural direct effects (NDE) for all mediators identified were not found to be significant, implying that the ITT effect appears to be operating primarily through the specified mediator (i.e., *omnipotence*, *assertiveness*, voice *acceptance* and *autonomous actions*).

### Adherence to treatment mediation (CACE)

Fifty-six (75%) of the 75 individuals randomised to AVATAR therapy met the adherence criteria i.e. had completed their avatar set-up and received at least three sessions of therapy, corresponding to the observed proportion of compliers in the treatment group. Overall, the complier specific mediated and direct effects were greater in magnitude than those from the ITT decomposition. For example, as can be seen for the voice omnipotence mediator, adjusting for baseline *omnipotence* and PSYRATS-AH-Total scores, a CACE analysis suggests that the average causal effect of receiving AVATAR therapy among the subgroup of individuals who comply with the intervention decreases PSYRATS-AH-Total score at week 12 by 4.51 points (SE = 1.96, 95% CI -8.32 to -0.87).

The CACE decomposition identified the same set of mediators as the ITT decomposition. Among the subgroup of individuals who would comply with the intervention, treatment receipt decreased PSYRATS-AH-Total score at week 12 via a reduction in the participants perceived *omnipotence* of the voice(s) by 1.69 points (SE = 1.15, 95% CI -4.36 to -0.06), while holding all other pathways constant. Similarly, as for the ITT results, the corresponding direct effects (CANDE) for all mediators were not found to be significant, implying that the CACE effect appears to be operating primarily through the specified mediator.

A sensitivity analysis identified 70 of the 75 (93%) individuals randomised to AVATAR therapy had at least completed their avatar set-up. Our primary mediation findings were robust to the choice of clinical cut point used to define adherence (see Table S1 in Supplementary material).

### ITT and CACE mediation of secondary outcomes

Similar findings were observed for the analyses with the other two outcomes variables PSYRATS-AH-Frequency and PSYRATS-AH-Distress outcome at 12 weeks (Please see Supplementary Materials S1 and S2) with some differences summarised below.

As with overall voice severity (PSYRATS-Total) *omnipotence*, *assertiveness* and *acceptance* were identified mediators for both ITT and CACE for PSYRATS-AH-Frequency and PSYRATS-AH-Distress, whereas malevolence, anxiety and self-esteem were not. Also, as was the case for overall voice severity, autonomous *action* in relation to voices (VAAS) was also mediator in both ITT and CACE for PSYRATS-AH-Distress but not for PSYRATS-AH-Frequency.

Lastly, for the PSYRATS-AH-Distress outcome, the corresponding direct effects were not found to be significant, implying that both the ITT and CACE effects appear to be operating primarily through the specified mediator. In contract, for the PSYRATS-AH-Frequency outcome at 12 weeks, corresponding direct effects were also statistically significant, suggesting that while *omnipotence*, *acceptance*, and *assertiveness* contributed to improvements in this outcome, additional pathways not captured in the model may also be contributing to the overall treatment effect.

**Table 2:**
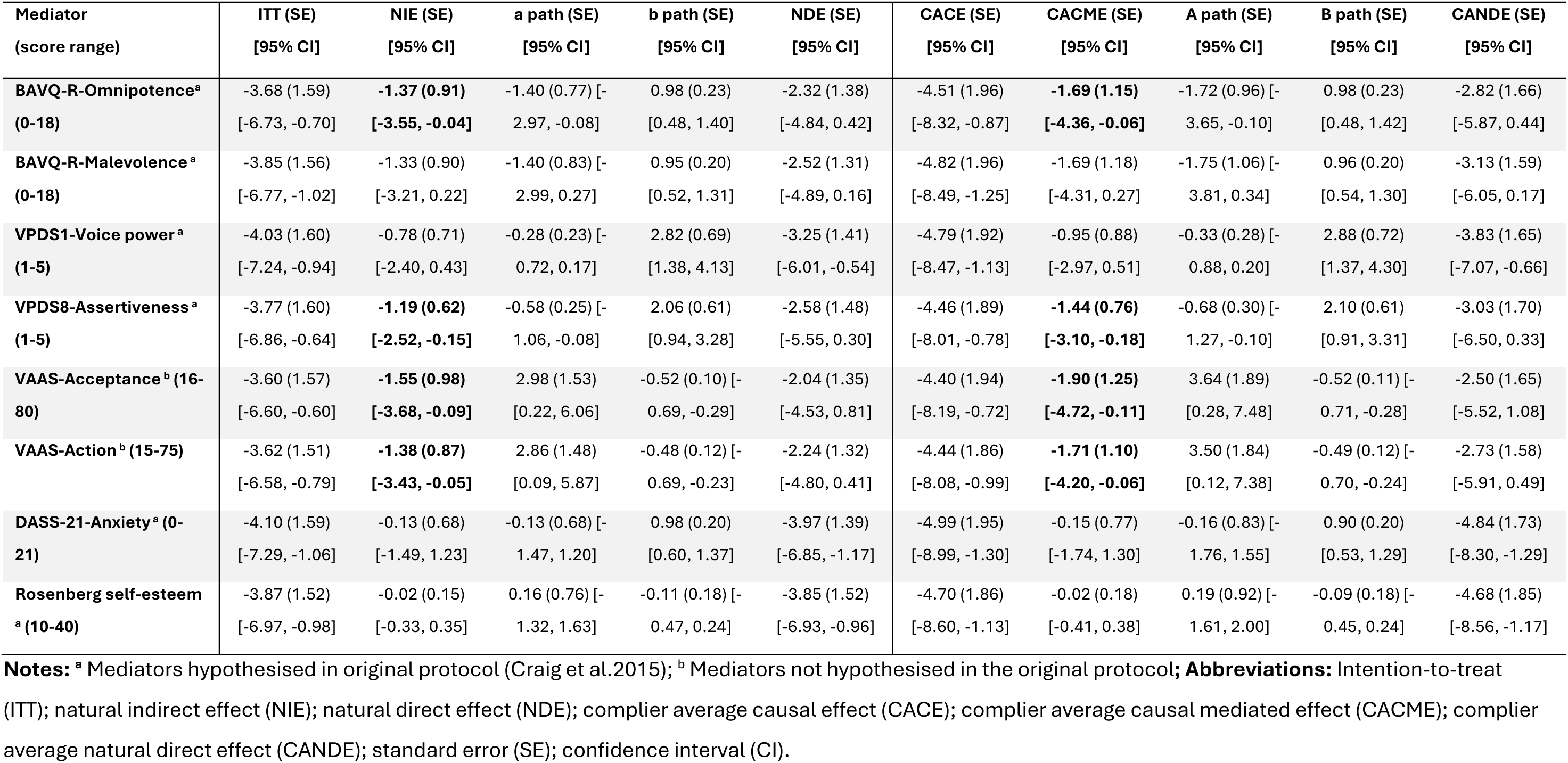
Estimates of the ITT and its decomposition into NIE and NDE, and of the CACE and its decomposition into CACME and CANDE, for the **PSYRATS-AH-Total score** at 12 weeks, with bootstrap standard errors and 95% confidence intervals (percentile bootstrap).

| Mediator<br>(score range) | ITT (SE)<br>[95% CI] | NIE (SE)<br>[95% CI] | a path (SE)<br>[95% CI] | b path (SE)<br>[95% CI] | NDE (SE)<br>[95% CI] | CACE (SE)<br>[95% CI] | CACME (SE)<br>[95% CI] | A path (SE)<br>[95% CI] | B path (SE)<br>[95% CI] | CANDE (SE)<br>[95% CI] |
| --- | --- | --- | --- | --- | --- | --- | --- | --- | --- | --- |
| **BAVQ-R-Omnipotence<sup>a</sup>**<br>**(0-18)** | -3.68 (1.59)<br>[-6.73, -0.70] | **-1.37 (0.91)**<br>**[-3.55, -0.04]** | -1.40 (0.77) [-<br>2.97, -0.08] | 0.98 (0.23)<br>[0.48, 1.40] | -2.32 (1.38)<br>[-4.84, 0.42] | -4.51 (1.96)<br>[-8.32, -0.87] | **-1.69 (1.15)**<br>**[-4.36, -0.06]** | -1.72 (0.96) [-<br>3.65, -0.10] | 0.98 (0.23)<br>[0.48, 1.42] | -2.82 (1.66)<br>[-5.87, 0.44] |
| **BAVQ-R-Malevolence<sup>a</sup>**<br>**(0-18)** | -3.85 (1.56)<br>[-6.77, -1.02] | -1.33 (0.90)<br>[-3.21, 0.22] | -1.40 (0.83) [-<br>2.99, 0.27] | 0.95 (0.20)<br>[0.52, 1.31] | -2.52 (1.31)<br>[-4.89, 0.16] | -4.82 (1.96)<br>[-8.49, -1.25] | -1.69 (1.18)<br>[-4.31, 0.27] | -1.75 (1.06) [-<br>3.81, 0.34] | 0.96 (0.20)<br>[0.54, 1.30] | -3.13 (1.59)<br>[-6.05, 0.17] |
| **VPDS1-Voice power<sup>a</sup>**<br>**(1-5)** | -4.03 (1.60)<br>[-7.24, -0.94] | -0.78 (0.71)<br>[-2.40, 0.43] | -0.28 (0.23) [-<br>0.72, 0.17] | 2.82 (0.69)<br>[1.38, 4.13] | -3.25 (1.41)<br>[-6.01, -0.54] | -4.79 (1.92)<br>[-8.47, -1.13] | -0.95 (0.88)<br>[-2.97, 0.51] | -0.33 (0.28) [-<br>0.88, 0.20] | 2.88 (0.72)<br>[1.37, 4.30] | -3.83 (1.65)<br>[-7.07, -0.66] |
| **VPDS8-Assertiveness<sup>a</sup>**<br>**(1-5)** | -3.77 (1.60)<br>[-6.86, -0.64] | **-1.19 (0.62)**<br>**[-2.52, -0.15]** | -0.58 (0.25) [-<br>1.06, -0.08] | 2.06 (0.61)<br>[0.94, 3.28] | -2.58 (1.48)<br>[-5.55, 0.30] | -4.46 (1.89)<br>[-8.01, -0.78] | **-1.44 (0.76)**<br>**[-3.10, -0.18]** | -0.68 (0.30) [-<br>1.27, -0.10] | 2.10 (0.61)<br>[0.91, 3.31] | -3.03 (1.70)<br>[-6.50, 0.33] |
| **VAAS-Acceptance<sup>b</sup> (16-80)** | -3.60 (1.57)<br>[-6.60, -0.60] | **-1.55 (0.98)**<br>**[-3.68, -0.09]** | 2.98 (1.53)<br>[0.22, 6.06] | -0.52 (0.10) [-<br>0.69, -0.29] | -2.04 (1.35)<br>[-4.53, 0.81] | -4.40 (1.94)<br>[-8.19, -0.72] | **-1.90 (1.25)**<br>**[-4.72, -0.11]** | 3.64 (1.89)<br>[0.28, 7.48] | -0.52 (0.11) [-<br>0.71, -0.28] | -2.50 (1.65)<br>[-5.52, 1.08] |
| **VAAS-Action<sup>b</sup> (15-75)** | -3.62 (1.51)<br>[-6.58, -0.79] | **-1.38 (0.87)**<br>**[-3.43, -0.05]** | 2.86 (1.48)<br>[0.09, 5.87] | -0.48 (0.12) [-<br>0.69, -0.23] | -2.24 (1.32)<br>[-4.80, 0.41] | -4.44 (1.86)<br>[-8.08, -0.99] | **-1.71 (1.10)**<br>**[-4.20, -0.06]** | 3.50 (1.84)<br>[0.12, 7.38] | -0.49 (0.12) [-<br>0.70, -0.24] | -2.73 (1.58)<br>[-5.91, 0.49] |
| **DASS-21-Anxiety<sup>a</sup> (0-21)** | -4.10 (1.59)<br>[-7.29, -1.06] | -0.13 (0.68)<br>[-1.49, 1.23] | -0.13 (0.68) [-<br>1.47, 1.20] | 0.98 (0.20)<br>[0.60, 1.37] | -3.97 (1.39)<br>[-6.85, -1.17] | -4.99 (1.95)<br>[-8.99, -1.30] | -0.15 (0.77)<br>[-1.74, 1.30] | -0.16 (0.83) [-<br>1.76, 1.55] | 0.90 (0.20)<br>[0.53, 1.29] | -4.84 (1.73)<br>[-8.30, -1.29] |
| **Rosenberg self-esteem<sup>a</sup> (10-40)** | -3.87 (1.52)<br>[-6.97, -0.98] | -0.02 (0.15)<br>[-0.33, 0.35] | 0.16 (0.76) [-<br>1.32, 1.63] | -0.11 (0.18) [-<br>0.47, 0.24] | -3.85 (1.52)<br>[-6.93, -0.96] | -4.70 (1.86)<br>[-8.60, -1.13] | -0.02 (0.18)<br>[-0.41, 0.38] | 0.19 (0.92) [-<br>1.61, 2.00] | -0.09 (0.18) [-<br>0.45, 0.24] | -4.68 (1.85)<br>[-8.56, -1.17] |

**Notes:** <sup>a</sup> Mediators hypothesised in original protocol (Craig et al.2015); <sup>b</sup> Mediators not hypothesised in the original protocol; **Abbreviations:** Intention-to-treat (ITT); natural indirect effect (NIE); natural direct effect (NDE); complier average causal effect (CACE); complier average causal mediated effect (CACME); complier average natural direct effect (CANDE); standard error (SE); confidence interval (CI).

## Discussion

The present study was the first mediation analysis of AVATAR therapy examining the role of power-related constructs. We investigated the psychological mechanisms through which treatment effects for AVATAR therapy are demonstrated on distressing voices. Specifically, we examined whether treatment effects on total severity, frequency, and distress were mediated by changes in beliefs about voices, self-esteem, appraisal of the voice relationship, and anxiety, as hypothesised in the AVATAR1 trial protocol. We also explored voice acceptance and action as additional potential mediators, and examined mediation effects using both ITT and CACE approaches to distinguish between treatment offer and treatment receipt effects.

As hypothesised, across both ITT and CACE analyses, reductions in perceived voice omnipotence and increased assertiveness in relation to voices, emerged as consistent mediators of AVATAR therapy on reductions in overall severity, frequency and distress of auditory hallucinations compared to SC, whereas voice malevolence, power differential, self-esteem and anxiety did not. Exploratory analysis also revealed that increases in acceptance and autonomous actions in relation to voices mediated the impact of AVATAR therapy on overall voice severity and distress. These findings are notable given that these processes are central to theoretical models underpinning AVATAR therapy [6-10, 14-16].

### Omnipotence, malevolence and power of voices

Among the identified mediators, perceived omnipotence of voices showed one of the most consistent mediation effects across main clinical outcomes. Participants receiving AVATAR therapy reported reductions in the extent to which voices were experienced as powerful, dominant, and controlling, which in turn was associated with reductions in voice severity. This finding aligns closely with cognitive models of voice-hearing, which propose that beliefs about the authority of voices play a central role in maintaining distress and submissive responding. AVATAR therapy may target these beliefs by allowing participants to directly confront and challenge the avatar representing the voice, without the expected retaliation from the voice or the anticipated negative consequences, potentially weakening perceptions of the voice as all-dominant (or directly “omnipotent”).

The broader construct of omnipotence appeared to capture the mechanisms of change in AVATAR therapy more accurately than the relative power between voice and voice hearer, which did not mediate improvements on outcomes. This echoes findings from COMMAND [55], which similarly reported that voice power differential alone did not mediate the impact of cognitive therapy on reducing harmful compliance with voices, whereas the broader scale (covering superiority, knowledge, and ability to inflict harm) did.

Perceived voice malevolence did not emerge as significant mediator of treatment effects, neither did it show significant improvement at the end of therapy [36], despite evidence that persecutory attributions towards the avatar decreased over the course of treatment [31].

Whereas malevolence reflects perceptions of hostile intent, omnipotence captures the voice’s perceived power, authority, and capacity to exert control over the individual [11]. Reducing the perceived power of voices may be more proximal to therapeutic change in AVATAR than modifying beliefs about their harmful intent, with participants developing greater assertiveness and agency even when the content remains critical or threatening [33].

### Assertiveness, acceptance and self-esteem

The use of experiential dialogue with voice representation is a core therapeutic technique of the new wave of relational therapies for voices [18]. AVATAR therapy primarily targeted the interpersonal dynamic between person and voice — specifically experiences of submission and diminished agency. Accordingly, as well as decreases in voice omnipotence appraisals, voice hearers reported behavioural changes of increased assertiveness which mediated outcomes in AVATAR therapy, whereas evaluative beliefs about the self (self-esteem) did not emerge as a significant mediator of treatment effects.

Voice acceptance, an unexpected mediator of AVATAR therapy across all outcomes in the present paper, warrants further attention. Acceptance of voice-hearing experiences increased during AVATAR therapy and mediated improvements in voice related outcomes. It is unclear how voice acceptance relates to reconciling with voices by ‘*gaining insights and experiencing revelations about the voices’* as voice hearers put it when describing the impact of Talking with Voices, another type of relational therapy [56]. Given previous associations between voice acceptance and reduced resistance to command hallucinations [46, 57], future research should examine how acceptance processes interact with behavioural changes, particularly those related to assertiveness and autonomy, as well as meaning making.

These findings are consistent with previous studies using qualitative [26-28] and observational research [29] documenting increases in assertive and reductions in submissive responses during avatar dialogues, as well as participants’ reports of feeling more empowered to stand up to their avatar.

### Anxiety and habituation to distressing voice content

Anxiety has long been considered a central factor in maintaining distressing voices [58]. Accordingly, we had hypothesised that habituation resulting from repeated exposure to distressing voice content and imagery would represent one of the key mechanisms of action in AVATAR therapy [34]. However, this hypothesis was not supported by the present findings. One possible explanation is that the measure used may not have captured the aspects of anxiety most relevant to voice-hearing experiences. Specifically, the Depression Anxiety Stress Scales (DASS) [48] primarily assesses general anxiety symptoms (e.g., “I was worried about situations in which I might panic and make a fool of myself”) rather than anxiety specifically related to hearing voices.

Interestingly, the recent AVATAR2 trial evaluating an extended version of AVATAR therapy [59] found significant post-treatment reductions in anxiety, as measured by the DASS-21 compared to treatment-as-usual’ (TAU) control. However, findings from Experience Sampling Methodology (ESM) data from the same trial showed mixed findings in relation to voice-related anxiety in everyday life [33]. Existing research consistently shows that participants experience avatar dialogues as anxiety-provoking [27, 31], and this specific anxiety during dialogues reduces across AVATAR therapy sessions [31]. Taken together, these findings suggest that AVATAR therapy may successfully reduce anxiety within the therapeutic context, but further research is needed to determine the extent to which these changes generalise to everyday voice-hearing experiences.

### Adherence to AVATAR therapy

As expected, the CACE analyses broadly replicated the ITT findings, with larger mediated and overall effects observed among participants who adhered to treatment. This strengthens confidence that the identified mechanisms reflect active engagement with AVATAR therapy, rather than treatment allocation alone. The larger effects among compliers suggest that repeated experiential engagement with the avatar dialogue helps to produce meaningful shifts in voice-related beliefs and interpersonal responding. This could be associated with therapy and voice hearer factors linked to engagement and participation, previous research has shown that participants who experience highly characterised voices engage in longer dialogues with their avatars during therapy [60].

### Strengths and limitations

This was the first study to evaluate power-related constructs mediators of outcomes in AVATAR therapy. The active trial design enabled mediated effects to be attributed to the specific therapeutic value of AVATAR therapy over and above non-specific factors, and both ITT and treatment adherence-based approaches were examined whilst controlling for baseline variables.

Several limitations warrant consideration. Confidence intervals for the indirect effects in both the ITT and CACE analyses were close to 0. This could be due to low magnitude of the indirect effect, but it is also likely that this reflects insufficient power given that the sample size was calculated to detect treatment effects on the primary outcome rather than mediated effects.

The ITT and CACE decompositions require a conditionally ignorable observed mediator (among compliers, for the CACE) for identification of effects. Whilst we adjusted for baseline mediator and outcome scores to increase the plausibility of this assumption, the relationship between the mediator and outcome may be subject to unmeasured, or post-randomisation confounding. As a single-blind study, participants’ awareness of their allocated treatment group may have influenced their behaviour, limiting the plausibility of the exclusion restriction assumption [61]. Additionally, the dichotomisation of the continuous treatment receipt variable (number of sessions attended) may further undermine the assumption’s reliability. Our findings are robust to the choice of cut point used in our primary analysis: a sensitivity analysis using the cut point that most optimally supports the exclusion restriction produced consistent results.

While mediation analysis generally requires temporal ordering of the mediator and outcome, the causal effect of the mediator on the outcome, or the mechanistic effect, is inferred not through time-separation but based on the theoretical understanding of the mechanism process. Finally, our findings are valid provided missing data are Missing-At-Random (MAR), conditional on the covariates in the analysis model. This assumption cannot be formally tested using the observed data and is more plausible for the CACE analyses than for the ITT analyses when missingness is related to non-adherence.

### Clinical implications and future research

A range of AVATAR therapy developments are currently being investigated that will help further understand mechanisms of change. A large multicentre RCT investigating AVATAR therapy optimisation in comparison to TAU (AVATAR2; [59]) included a mediation analysis plan in their protocol with hypotheses on reductions in omnipotence and malevolence (BAVQ-R) as well as anxiety in daily life as measured by ESM. Technological innovations such as immersive virtual reality components in Canada [62] and Denmark [63] and more recently in planned work in Germany including social environments are currently being evaluated [64] and will help to quantify the impact of technological malleable factors such as sense of presence and enhance generalisability to everyday life.

Further research should consider the role of moderators on mediation. As ongoing social adversity is known to shape hallucinatory experiences [9], AVATAR therapy has explicitly aimed to acknowledge societal discrimination in minoritised communities and considered avatar dialogues as opportunities to rescript disempowering encounters [65]. It is an unknown if AVATAR therapy is as effective in voice hearers’ with current experiences of marginalisation and oppression.

The current mechanistic study showed that through interacting with an embodied representation of the voice, AVATAR therapy improves overall severity, frequency and distress of voices compared to SC, and that it does so by reducing their perceived omnipotence, promoting their acceptance and increasing voice hearers’ assertiveness and sense of agency in relation to voices. Identified mediators could be monitored during AVATAR therapy to ensure they are being effectively targeted.

## Supporting information

Supplementary materials_AVATAR1 mediation paper

## Data Availability

The data and code for the mediation analyses are available upon reasonable request.

## Acknowledgements

The trial was funded by Wellcome Trust (FWBC-AVATAR 098272/z/12/z). TW and AH are part-funded by the National Institute for Health and Care Research (NIHR) Biomedical Research Centre (BRC): Maudsley. The study was supported by the National Institute for Health and Care Research (NIHR), the NIHR Maudsley Biomedical Research Centre and Maudsley NHS Foundation Trust, King’s College London (PG; NIHR203318). The views expressed are those of the authors and not necessarily those of the NIHR or the Department of Health and Social Care. MRC discloses support from the Sofja Kovaleskaja Award, from the Alexander von Humboldt Foundation and Ministry of Education and Research (Germany, 3.2-1210962-GBR-SKP). The funding bodies did not have any role in study design; data collection, analysis, and interpretation; or writing of the manuscript. RE is funded by an NIHR Research Professorship (NIHR300051). CE discloses support from a Wellcome Early Career Award (227646/Z/23/Z). The funding bodies did not have any role in study design, data collection and analysis, data interpretation, or writing of the manuscript. We thank the trial participants who consented to take part and generously provided their data, and the clinical teams who referred and supported the participants

## Conflicts of interest

M.H. is a shareholder of Avatar Therapy Ltd. T.K.J.C. and P.A.G. are unpaid scientific advisers to Avatar Therapy Ltd. TW, MRC, TC and AH are co-founders of AVATAR therapy. The other authors declare no competing interests.

## Data sharing statement

The data and code for the mediation analyses are available upon reasonable request.

## Supplementary material

### S1 Secondary outcome (Voice Frequency)

Estimates of the ITT and its decomposition into NIE and NDE, and of the CACE and its decomposition into CACME and CANDE, for the PSYRAT-AH-Frequency at 12 weeks, with bootstrap standard errors and 95% confidence intervals (percentile bootstrap).

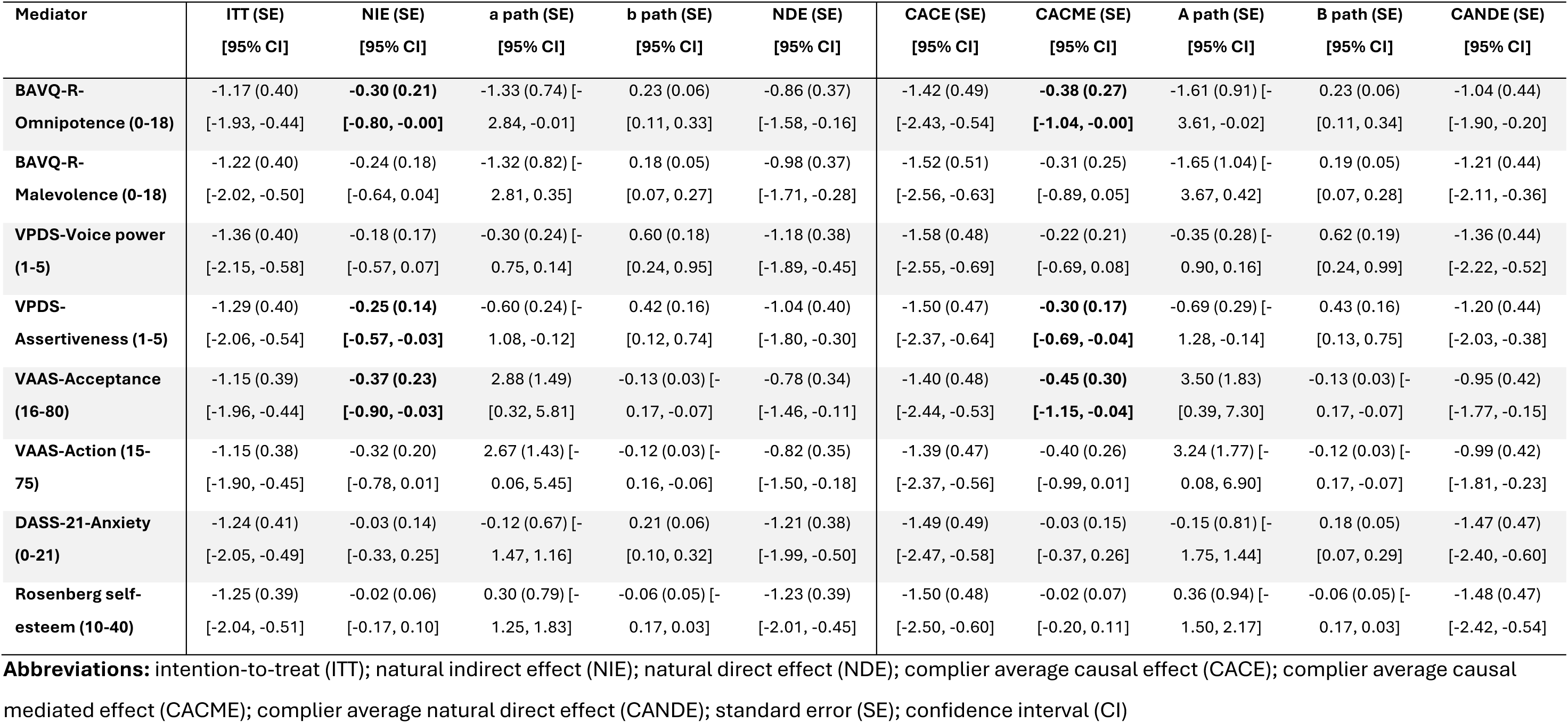

### S2 Secondary outcome (Voice Distress)

Estimates of the ITT and its decomposition into NIE and NDE, and of the CACE and its decomposition into CACME and CANDE, for the PSYRAT-AH-Distress at 12 weeks, with bootstrap standard errors and 95% confidence intervals (percentile bootstrap).

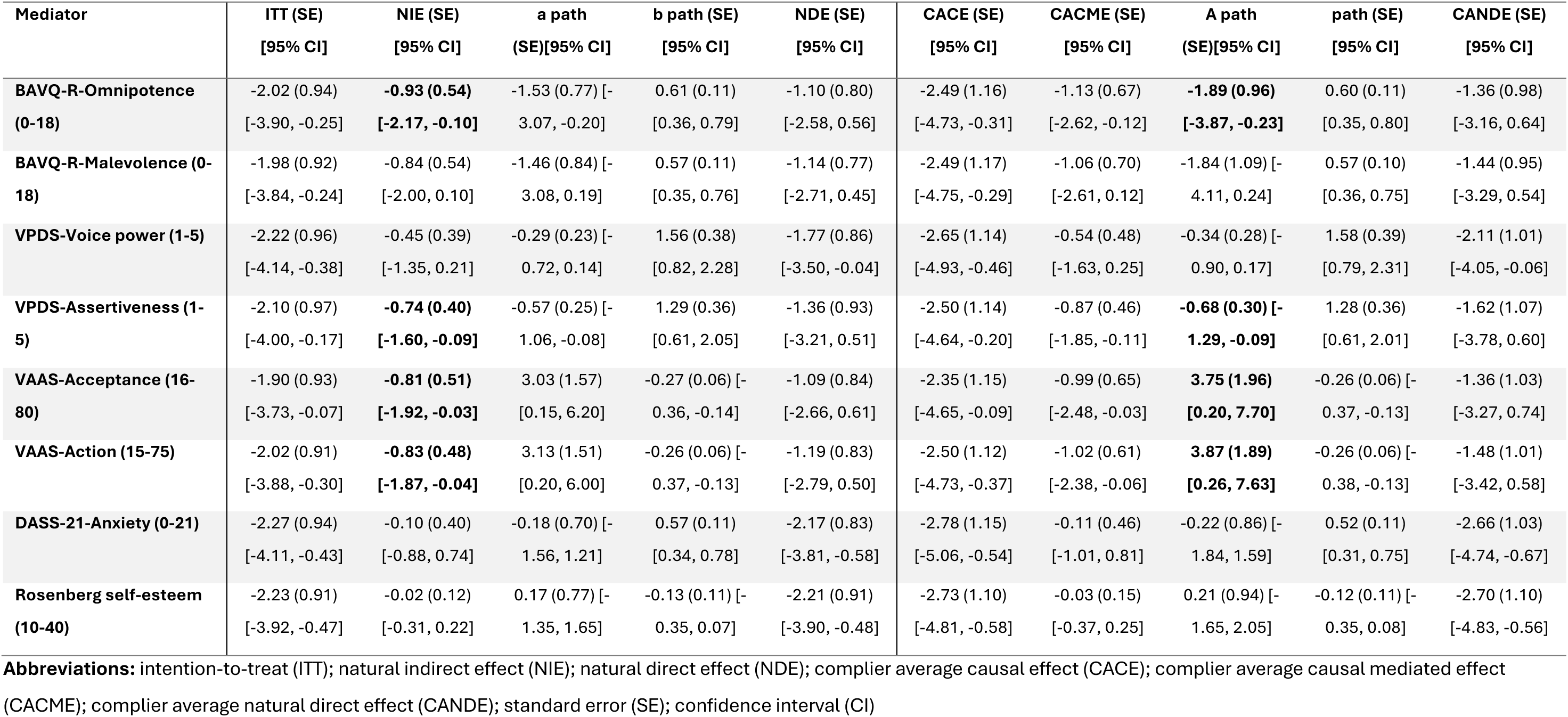

### S3 Sensitivity Analysis

Estimates of the ITT and its decomposition into NIE and NDE, and of the CACE and its decomposition into CACME and CANDE, for the **PSYRAT-AH-Total score** at 12 weeks, with bootstrap standard errors and 95% confidence intervals (percentile bootstrap).

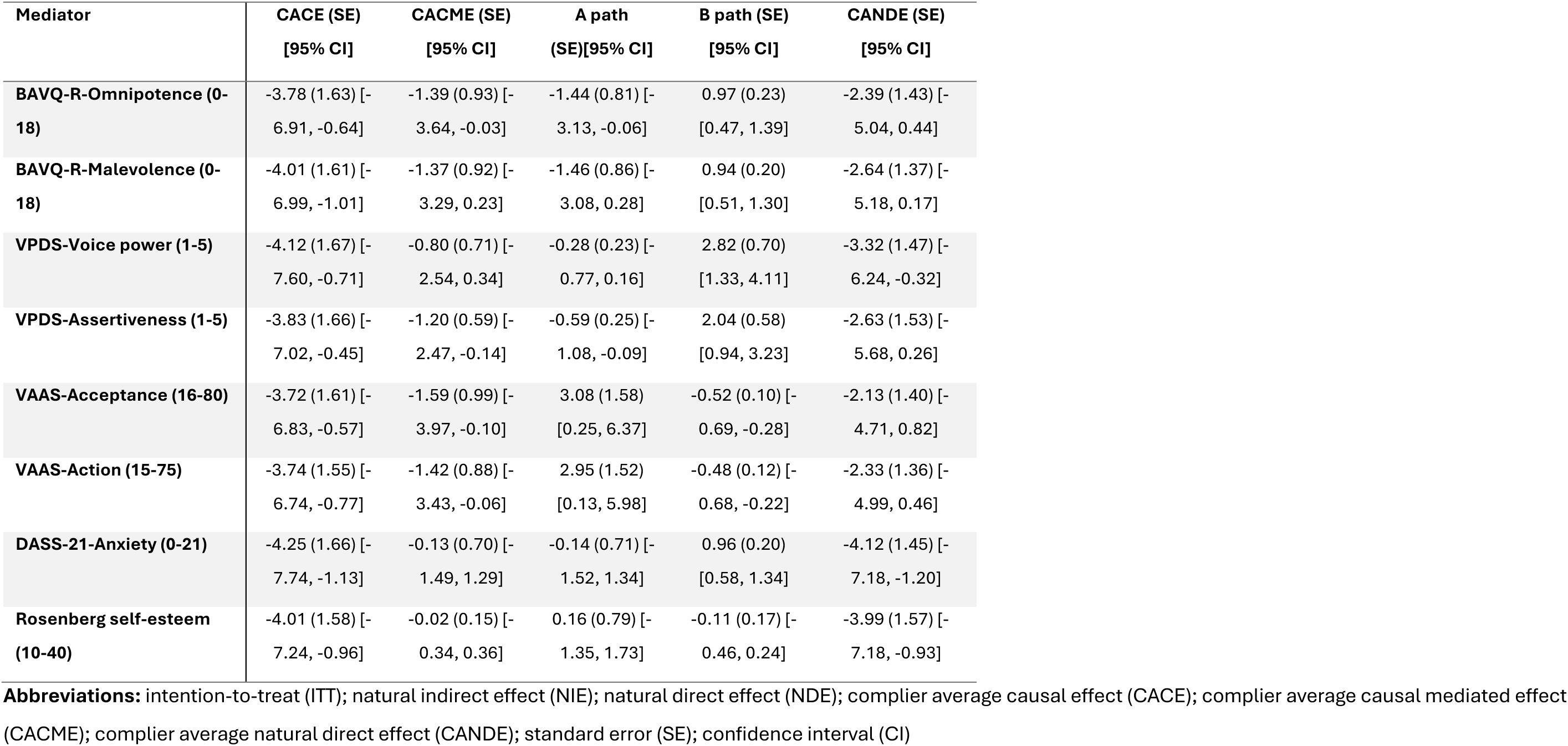

### S4 AGReMA Checklist

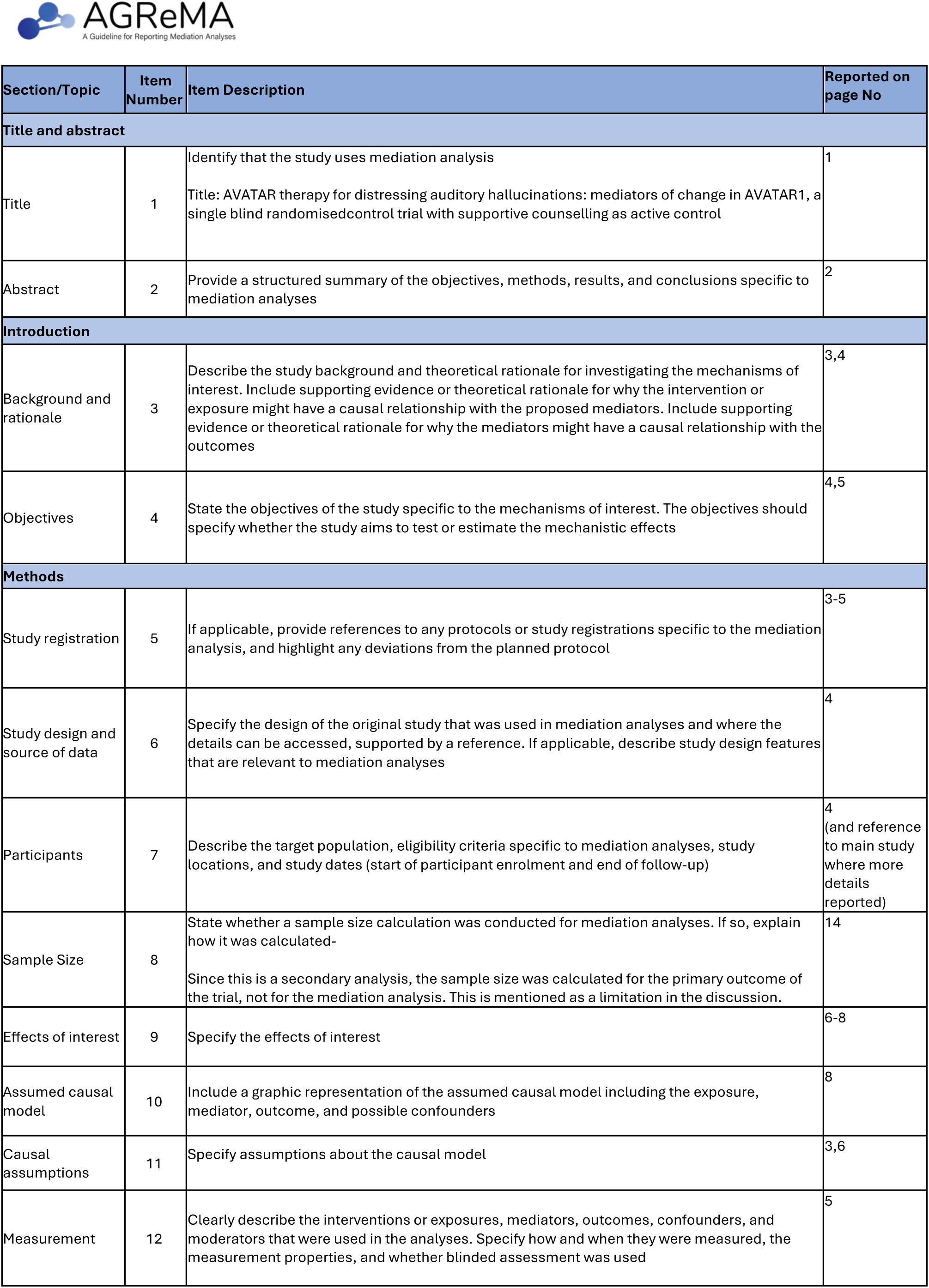
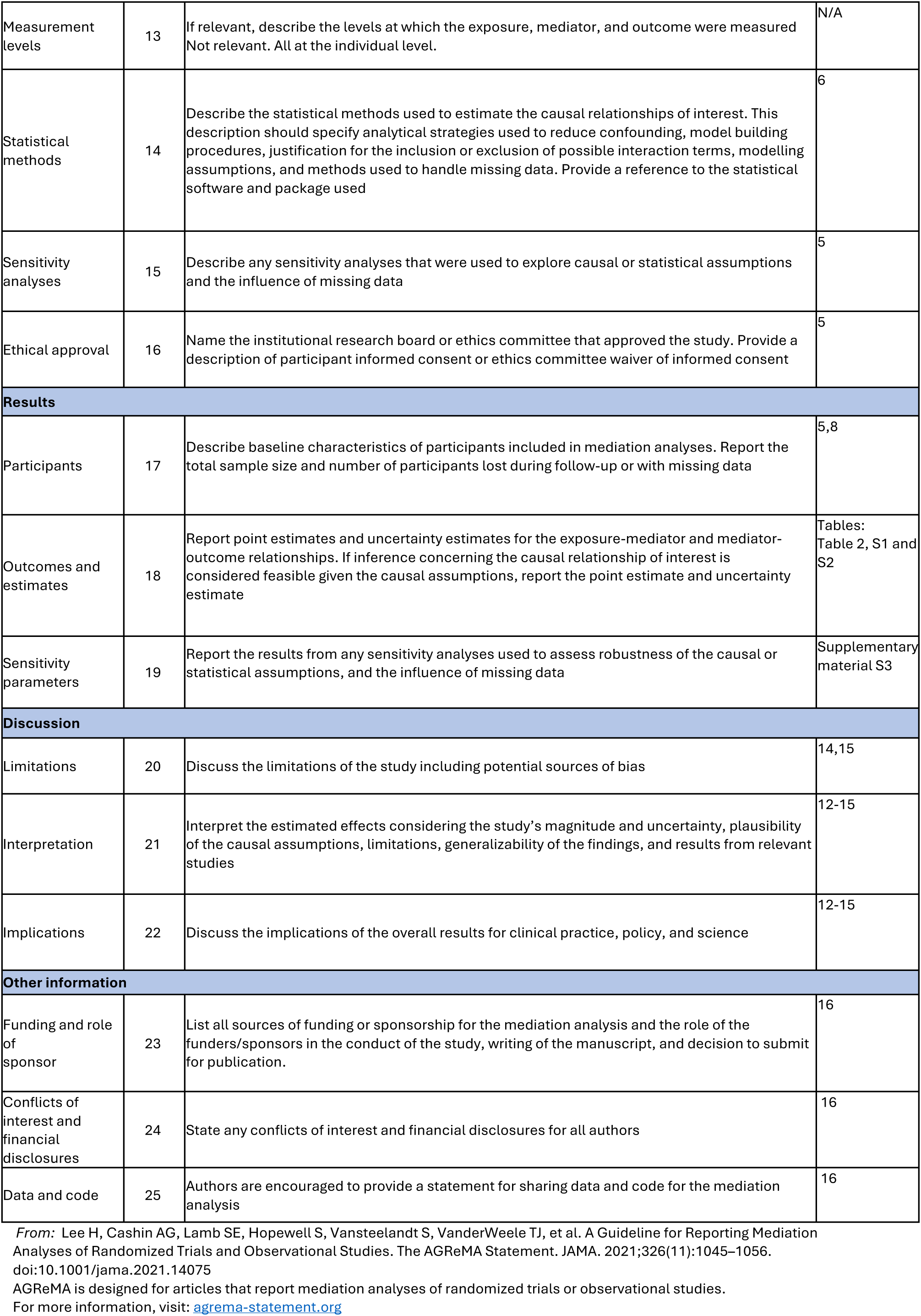

## Footnotes

1 Although most models acknowledge the presence of other factors, we have here listed the factor(s) that are the main focus emphasised by each of the models/ theories.

2 Please note Craig et al (2018) was initially named simply AVATAR trial. As there have been a subsequent recently published AVATAR2 trial (2024), which will include its own planned mediation this has been retrospectively labelled AVATAR1.

## Notes

### Clinical Trial

ISRCTN65314790

### Clinical Protocols

https://link.springer.com/article/10.1186/s13063-015-0888-6

### Author Declarations

The trial received ethical approved from the London-Hampstead Research Ethics Committee (reference 13/Lo/0482).

### Summary of Updates

We have ammended the following information: 1. We have added an additional author affiliations for Mar Rus-Calafell: j German Center for Mental Health (DZPG), Partner Site Bochum/Marburg, Bochum, Germany 2. In the conflicts of interest section, we have added authors MRC and MH to the following statement: TW, MRC, TC and AH are co-founders of AVATAR therapy.

